# Early brain changes in Lyme disease are associated with clinical outcomes

**DOI:** 10.1101/2024.12.16.24319088

**Authors:** Cherie L. Marvel, Alison W. Rebman, Kylie H. Alm, Pegah Touradji, Arnold Bakker, Prianca A. Nadkarni, Deeya Bhattacharya, Owen P. Morgan, Amy Mistri, Christopher C. Sandino, Jonathan A. Ecker, Erica A. Kozero, Arun Venkatesan, Abhay Moghekar, Ashar A. Keeys, John N. Aucott

## Abstract

In Lyme disease (LD), 10-20% of patients develop persistent symptoms following antibiotic treatment (i.e., post-treatment Lyme disease (PTLD)), which often includes neurological symptoms. This study tested the hypothesis that brain changes occur early in LD and influence outcomes.

Functional MRI (fMRI) of working memory and health surveys were administered to people with acute LD (n=20) after treatment and six months later. Demographically matched healthy controls (HC; n=19) were also assessed six months apart. The LD group was categorized at six months into those who had returned to health (RTH, n=11) or reported persistent symptoms (sPTLD, n=9). FMRI data from both LD subgroups were compared to each other and HC at both time points. Brain regions of interest (ROI) values were compared to health surveys.

Baseline brain activity in RTH was elevated in fronto-parietal regions relative to sPTLD (p < .0025) and HC (p < .001). Notably, 64% of activation clusters in RTH were in white matter, confirmed by segmentation analysis. ROI values from RTH vs. HC correlated with higher health survey scores. At 6-months, few group differences remained. By contrast, ROI values from sPTLD vs. HC yielded few activation differences at either time point or correlations with health survey scores.

Robust brain activity in early LD was associated with future RTH. By contrast, the absence of early brain activity was associated with persistent symptoms, suggesting that failure to mount an early response contributes to PTLD. Understanding how brain activity relates to recovery in LD can aid prognosis and guide treatment.

## Introduction

Lyme disease (LD) is an inflammatory disease initiated by infection with *Borrelia burgdorferi* (Bb) following the bite of an infected tick. After standard of care antibiotic therapy, 10-20% of patients treated for LD will develop a chronic syndrome of patient-reported symptoms, known as post-treatment Lyme disease (PTLD). [1, 2] The Infectious Disease Society of America (IDSA)-proposed case definition of PTLD includes prior physician-documented LD, appropriate antibiotic treatment, and development of subjective complaints of fatigue, widespread musculoskeletal pain, and/or cognitive difficulties within six months of the LD diagnosis that lasts for at least six months, and results in significant social or functional decline. [3–5] Little is known about the pathophysiology of or risk factors for PTLD, and there are no FDA-approved therapeutics for curative treatment. Importantly, clinicians currently cannot identify patients who are at risk of developing PTLD at the time of acute infection. [6]

Patient-reported cognitive symptoms are common among patients with PTLD, with complaints as high as 92% of those interviewed. [6, 7] However, cognitive impairments can be associated with other illness-related factors, such as sleep disturbance, mood, fatigue, pain, and poor health-related quality of life, all of which can be significant in those with PTLD. [5] To date, few investigations *in vivo* brain studies of LD patients have been conducted, noting glial cell activity and abnormal cerebral blood flow. [8–10] Recently, PTLD and healthy control participants performed a cognitive task while undergoing functional MRI (fMRI). [11] In the PTLD group relative to controls, white matter activations were observed, a phenomenon which is highly unusual. [12–14] Moreover, imaging characteristics within these white matter activations correlated with *<u>fewer</u>* cognitive and neurological symptoms. Thus, evidence suggests that the brain can be altered by LD, these alterations involve white matter, and, perhaps counterintuitively, such changes are associated with better clinical outcomes.

The current case-control study examined brain changes associated with early LD within weeks of the first diagnosis of acute infection and again six months later to capture the onset of white matter changes in LD. We hypothesized that white matter changes would emerge within the first six months post-infection and would be associated with favorable clinical outcomes, consistent with prior findings in a cross-sectional PTLD cohort. [11]

## Materials and methods

### Participants

Adults with LD were recruited from the Johns Hopkins Lyme Disease Research Center in which they were enrolled in the on-going <u>S</u>tudy of <u>L</u>yme <u>I</u>mmunology and <u>C</u>linical <u>E</u>vents (SLICE). [15] For SLICE enrollment, participants were required to have a visible, diagnostic erythema migrans rash ≥ 5 cm and to not have been ill longer than 3 months. Patients were excluded if they self-reported a medical history of any conditions with significant symptom overlap with PTLD, similar to those listed in the IDSA’s proposed case definition. [3] Specifically, those with fibromyalgia, chronic fatigue, major immunosuppression, psychiatric or autoimmune illnesses, hepatitis B/C, HIV, cancer chemotherapy treatment in the past two years, a history of illicit drug or substance abuse, Long COVID, or current pregnancy were excluded.

Healthy controls (HC) were recruited via community flyers and screened for any co-morbid conditions with significant symptom overlap with PTLD, as well as the exclusion criteria described above, or a past diagnosis of LD.

In a final screening stage, both LD and HC participants were excluded from study participation if they endorsed the following characteristics that might confound data interpretation: history of major neurologic disorders (including history of stroke, seizures, or HIV); a prior head injury resulting in loss of consciousness for longer than 5 minutes; current severe or unstable medical disorder; history before LD of a significant learning disability; and history before LD of a severe mood or psychotic disorder. Participants were also excluded for contraindications within the MRI environment, such as surgical implants and claustrophobia. Finally, participants were excluded for left-handedness or being a non-native English speaker (unless English was learned before puberty) because each of these would confound results due to the nature of the verbal working memory task administered during fMRI.

Participants were asked to return six months after their MRI to re-assess clinical and cognitive variables, obtain a second MRI, and determine health outcomes post infection. Data were removed from analyses if a participant dropped out of the SLICE study before an outcome could be ascertained, fMRI data was unusable, or there was an incidental MRI finding. In some cases, participants did not return for their follow-up MRI, but remained in the SLICE study, enabling a health outcome determination, and their baseline data were included.

The Institutional Review Board of the Johns Hopkins University School of Medicine approved this study. Written informed consent was obtained according to the Declaration of Helsinki from all participants prior to initiation of study activities, and all participants received compensation.

### Clinical assessment

Overall health, mood, symptom severity, fatigue, pain and health-related quality of life were assessed by standardized questionnaires which were administered at the baseline and 6-month visits: Short Form Health Survey, version 2 (SF-36) [16], Fatigue Severity Scale (FSS) [17], Short-Form McGill Pain Questionnaire-2 (SF-MPQ-2) [18], Beck Depression Inventory II (BDI-II) [19], and a self-administered a 36-item measure of symptom presence and severity (the post-Lyme Questionnaire of Symptoms [PLQS]). [20] For questionnaires, higher scores indicated greater symptom severity, except on the SF-36 where higher scores indicated higher health-related quality of life and functioning. (Online Table1)

Patient outcome status was determined at the 6-month follow-up visit. Due to the relatively small sample size, we dichotomized patients into two groups only; those who met criteria for return to health (RTH) vs. all others, who were considered sPTLD with either symptoms and/or functional impact present. [20, 21] Patients met criteria for RTH if: a) their PLQS did not indicate the presence of either fatigue, musculoskeletal pain, or cognitive complaints at the ‘moderate’ or ‘severe’ level, and b) their average composite score of 4 specific norm-based subscales on the SF-36 was no greater than 0.5 SD below the population mean. [21] Remaining patients not meeting criteria for RTH were considered sPTLD with either symptoms and/or functional impact present.

### Cognitive assessment

Standardized cognitive tests were administered at each timepoint. (see Online Table 2 for a list of tests) Raw scores were converted to T-scores using established norms.

### fMRI tasks

#### Verbal working memory task

Participants performed a verbal working memory task in the MRI scanner that has been described previously. [11, 22, 23] The paradigm involved two task conditions, each consisting of two stimulus conditions, resulting in four overall study conditions. Briefly, in the “control” condition, participants visually encoded one or two letters, and after a 4-6 second delay, they indicated by button press whether a probe letter matched a target letter. In the “forward” condition, participants again visually encoded one or two letters. During the delay period, however, they counted two alphabetical letters forward of each letter and held those new letters in mind. For example, if the letters were ‘‘f” and ‘‘q”, participants would hold ‘‘h” and ‘‘s” in mind. When the probe letter appeared, participants indicated whether the probe matched the newly derived target letters. Thus, there were four conditions total: 1) one-letter control, 2) two-letters control, 3) one-letter forward, and 4) two-letters forward. Additional parameter details are provided in Online Methods1. The measures of interest were accuracy and response time (RT) for each condition, with no minimal performance threshold for data inclusion. Due to a technical error, responses from one control participant were not recorded. This participant was not included in the behavioral analysis but remained in the fMRI analysis by including all trials.

#### Hemodynamic response function (HRF) tapping task

Individualized HRFs were obtained for convolutions during the event-related fMRI analyses to account for potential HRF differences across participants or study groups. This was obtained via a tapping task involving the right index finger. Tapping blocks lasted ∼30 seconds upon presentation of “tap” instructions, followed by “rest”, also lasting ∼ 30 seconds, for a total duration of 10 minutes. [11, 22, 24, 25]

### MRI data acquisition

All MRI data were acquired on a Philips 3 Tesla scanner using a 32-channel head coil. Equipment information is detailed in Online Methods2.

#### Structural MRI

A sagittal magnetization prepared gradient-echo (MPRAGE) sequence aligned to the anterior-posterior commissure (AC-PC) axis was used with the following parameters: repetition time (TR)/echo time (TE) = 7/3.3 ms; field of view = 240 mm x 240 mm; 170 slices; slice thickness 1.0 mm; 0 mm gap; flip angle = 8 degrees; voxel size = 0.75 mm x 0.75 mm. The total scan duration was 6 minutes.

#### Functional MRI

A T2-weighted gradient echo EPI pulse sequence was used with the following parameters: TR = 1000 ms; TE = 30 ms; flip angle = 61 degrees; in-plane resolution = 3.75 mm; slice thickness = 6 mm with a 1 mm gap; 20 oblique-axial slices; FOV = 21mm x 240 mm. To maximize whole-brain coverage to include the cerebellum and neocortex, images were acquired in the oblique-axial plane rotated 25 degrees clockwise with respect to the AC-PC line. The number of acquired volumes within each run ranged from 917 to 922 for the working memory tasks and 630 for the tapping task. The start of the fMRI scan was triggered by Eprime 2.0 software (Psychology Software Tools, Pittsburgh, PA) at the beginning of each run.

### MRI data analysis

#### Functional data analysis

Standard image preprocessing steps were performed using SPM12 [26]: slice timing correction (reference = slice #10), motion correction of all volumes aligned to the first volume of the first run, anatomical co-registration, normalization to the Montreal Neurological Institute (MNI) stereotaxic space, and spatial smoothing (FWHM = 8 mm). Individual HRF regressors were convolved with reference waveforms for the delay phase of the task (i.e., the 4-6 second delay between target and probe presentation) for each participant within the first-level analysis. This represented an event-related analysis focusing on working memory in the absence of visual stimuli or motor response. Statistical maps were computed for each participant using the general linear model approach with high pass filtering of 128 seconds. A random effects analysis was performed to map the average responses for correct trials only. All trials were included for one control at baseline whose behavioral data was not recorded due to a technical error. A beta contrast volume per participant was computed and used to conduct one-sample t-test values at every voxel. Within-group contrasts compared the blood oxygen level dependence (BOLD) signal difference between the two-letters forward minus two-letters control conditions. These differences were compared between group pairs (i.e., All Lyme vs. HC, RTH vs. HC, sPTLD vs. HC, and RTH vs. sPTLD). Activations were identified using a threshold of p < .001 with a cluster-level k ≥ 10. ROIs were created from surviving clusters using the Mars-BaR toolbox for SPM. [27] Individual ROI values were segmented for tissue classification. ROIs containing > 50% white matter were considered white matter activations. Gray and white matter activations were correlated with cognitive and clinical variables. For anatomical determinations of the activations, MNI coordinates were transformed into the coordinate system of the Talairach and Tourneaux stereotaxic atlas [28] using Bioimage Suite Web (Version 1.2.0) [29] and cross-referenced with atlas manuals. [28, 30]

#### Tissue class segmentation analysis

Following methods developed previously [11], significant fMRI activation clusters for each contrast and timepoint of interest were first transformed into binary ROIs masks, and then resampled into 1 mm^3^ space using nearest neighbor interpolation to match the MPRAGE template space used for tissue class segmentation. A study-specific T1 modal model template was generated using all participants and both timepoints. Advanced Normalization Tools (ANTs) software [31, 32] was used to calculate a 3D vector field transformation for each MPRAGE image input aligned to the study-specific template space. FSL’s FAST automated segmentation [33, 34] was used to segment the template into three tissue classes: gray matter, white matter, and cerebrospinal fluid. Each binary ROI mask was multiplied by the binary white matter segmentation mask, yielding the number of white matter voxels per ROI. This value was divided by the number of voxels in the entire ROI and multiplied by 100 to yield the percentage of white matter voxels. ROIs that contained > 50% white matter were categorized as “white matter activations” in subsequent analyses.

#### ROI analysis with clinical & cognitive variables

The estimate of activation within gray and white matter clusters were computed using Mars-BaR. [27] All voxel values within the ROI were averaged to yield one value per ROI. This value was correlated to variables obtained from questionnaires and cognitive tests.

### Statistical analysis

The clinical, cognitive, and MRI data contained continuous variables, except gender, which was categorical. In group comparisons (RTH vs. sPTLD, RTH vs. HC, and sPTLD vs. HC), t-tests and one-way ANOVA tests were used to compare continuous variables (e.g., age and education), and Fisher’s Exact test was used to compare gender. P-values for t-tests were determined by the result of the Levene’s Test for Equality of Variances. Shapiro-Wilk tests were used to determine if continuous variables followed a normal distribution. If a variable was not normally distributed, Mann-Whitney U-tests and Kruskal-Wallis tests were conducted to compare groups. Mixed-design ANOVAs were used to compare repeated measures between groups (e.g., fMRI task performance). [No ANOVAs contained a within-subjects factor with more than two levels; sphericity corrections were, therefore, not needed.] If mixed-design ANOVAs revealed group effects, pairwise t-tests were conducted to determine which groups were affected differentially. Pearson correlations were used when the Shapiro-Wilk’s normality tests indicated a normal data distribution (e.g., correlating ROI values with symptom measures). Spearman’s rho nonparametric correlations were used for variables with non-normal data distributions. However, ROI values were correlated to clinical and cognitive scores using Spearman’s correlations in all cases for a conservative approach in the absence of correction for multiple comparisons. Missing data within a symptom rating scale was imputed such that the mean for all other symptom items for an individual was entered as the missing value and fed into the summary score. All tests were two-tailed, with an alpha level < .05 to define statistical significance. Statistics were performed using IBM SPSS Statistics, Macintosh, version 29 (IBM Corp., Armonk, NY, USA).

## Results

### Participants

Adults with early LD (n=23) and HC (n=21) consented for this study between 2017-2022. A final sample of 20 LD participants were included in the baseline and 17 in the follow-up analysis; 19 HC participants were included in the baseline and 16 in the follow-up analysis (Fig. 1). LD participants completed their baseline fMRI visit with a mean of 15.2 days (range: 1-51) after completing doxycycline treatment and a mean of 42.3 days (range: 25-79) after the onset of their LD.

**Fig. 1.**
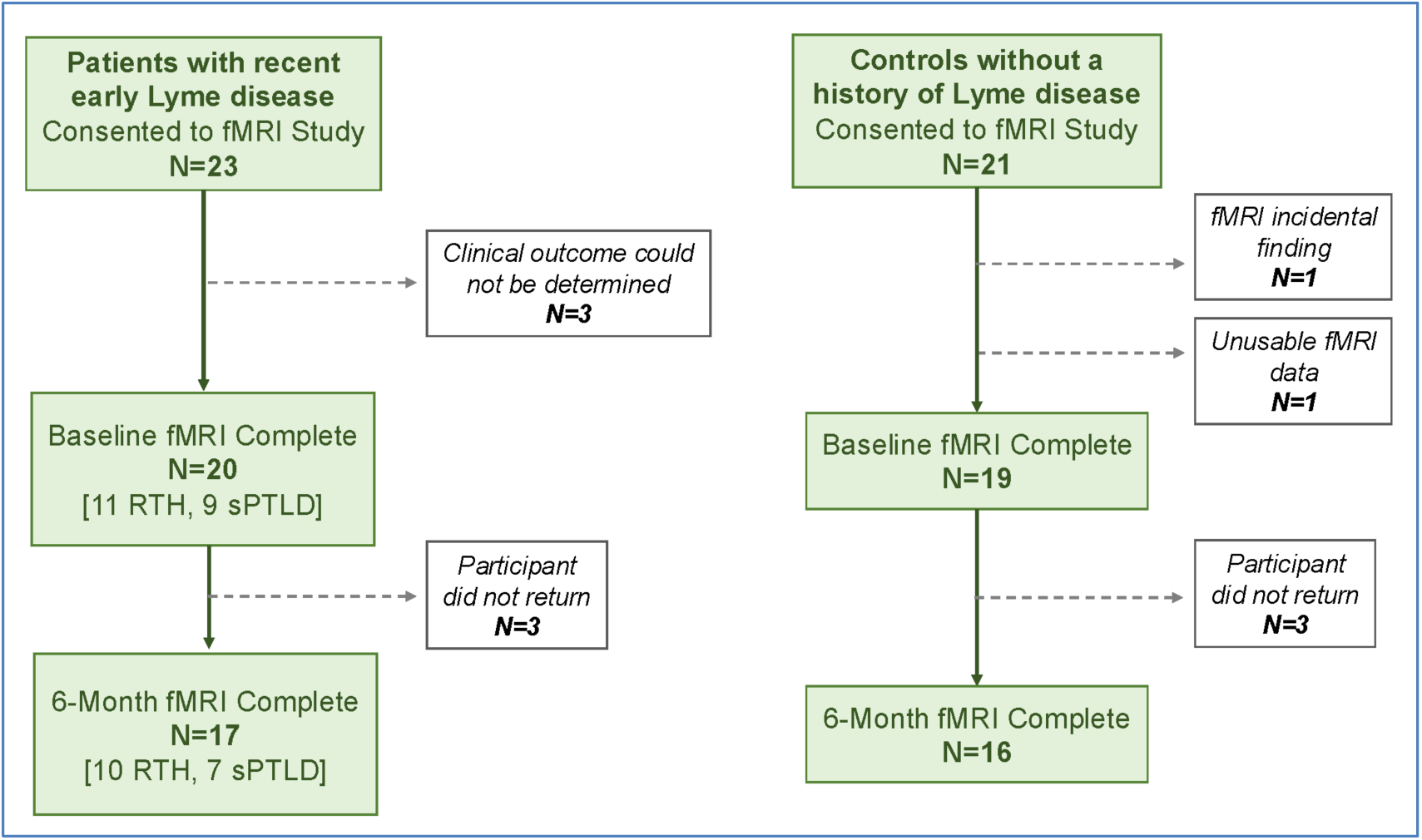
Study recruitment and retention data. Description of sample sizes for both study groups from consent to completion

Based on clinical and health measures obtained at six months, 11 Lyme participants were assigned to the RTH group and nine to the sPTLD group. (Table 1) Groups (sPTLD, RTH, and HC) did not differ by age *(H(2) = 3.02, p = .221*), years of education (*F*(2,36) *= .042, p = .959*), or gender, *p = .214*. Comparisons between the RTH and sPTLD groups revealed no differences in the median days’ duration between LD onset and receiving antibiotic treatment (RTH = 4.00 vs. sPTLD = 6.00, *U = 46.0, p = .819),* between LD onset and receiving the first MRI scan (RTH *=* 36.00 vs. sPTLD *=* 38.00, *U = 56.5, p = .603*), or between ending antibiotics and receiving the first MRI scan (RTH *=* 10.00 vs. sPTLD *=* 12.00, *U* = *56.5, p = .603*). Groups did not differ in the duration between MRI scans (*H(2) = .530, p = .767)*.

**Table 1.** Baseline demographic and clinical characteristics.

| Table 1 Baseline demographic and clinical characteristics |  |  |  |
| --- | --- | --- | --- |
|  | RTH (n=11) | sPTLD (n=9) | HC (n=19) |
| Age (years) | 56.51 (14.57) | 47.14 (14.72) | 51.16 [37.47, 57.93] |
| Male Gender | 5 (45.45%) | 6 (66.67%) | 6 (31.58%) |
| Education (Years) | 16.55 (2.62) | 16.78 (2.22) | 16.53 (1.98) |
| Two-Tier Seropositive<sup>1</sup> | 5/10<sup>1</sup> (50.00%) | 6 (66.67%) | 0 (0.00%) |
| Disseminated EM Lesions<sup>2</sup> | 6 (54.44%) | 3 (33.33%) | N/A |
| Lyme Disease Duration Prior to Treatment (Days) | 6.59 (4.85) | 6.00 [3.00, 6.00] | N/A |
| Lyme Disease Duration at first fMRI (Days) | 36.00 [31.00, 52.00] | 40.89 (8.49) | N/A |
| Days Since Stopping Doxycycline at First Scan (Days)<sup>3</sup> | 10.00 [4.00, 26.00] | 13.0 (4.97) | N/A |
| Days Between MRI Scans<sup>4</sup> | 192.00 (18.28), n=10 | 199.14 (23.57), n=7 | 184.50 [179.50, 201.50], n=16 |

N (%) are presented for categorical variables. Mean (standard deviation) are presented for normally distributed continuous variables, and median [IQR] are presented for non-normally distributed continuous variables. Sample sizes are presented when missing data occurred. No group differences were found across variables, all p-values > .214.
<sup>1</sup>Based on acute and convalescent testing interpreted according to CDC criteria which account for illness duration at the time of the test. One RTH patient was missing complete serostatus information.
<sup>2</sup>No patients had early disseminated neurologic or cardiac Lyme disease at the time of diagnosis.
<sup>3</sup>One person was at the end of their doxycycline treatment when the first scan was performed.
<sup>4</sup>One RTH, two sPTLD, and three HC participants did not return for their 6-month MRI scan.

### Clinical assessment

At baseline, the patient groups differed on the SF-36 (Social Functioning Scale, *U = 18.00, p = .033*; Mental Component Scale, *t*(17) *= 2.86, p = .011*) and the PLQS (Symptoms Total, *U = 68.50, p = .041*). At follow-up, patient groups differed on the BDI Total, *U = 11.00, p = .023*; the BDI Cognitive/Affective Subscale, *U = 13.00, p = .039*, and the PLQS (Symptoms Total, *U = 8.00, p = .003*). (Table 2)

**Table 2.** Standardized symptom measures.

| Table 2 Standardized symptom measures |  |  |  |  |
| --- | --- | --- | --- | --- |
|  | Baseline Visit |  | 6-Month Follow-up |  |
|  | RTH | sPTLD | RTH | sPTLD |

|  | *n*=11 | *n*=9 | *n*=10 | *n*=7 |
| --- | --- | --- | --- | --- |
| Beck Depression Inventory-II Total | 4.82 (3.76) | 9.43 (7.91), *n*=7 | .00 [.00, 3.00], *n*=9* | 6.57 (6.58)* |
| Beck Depression Inventory-II Cognitive/Affective Subscale | 1.00 [.00, 3.00] | 4.29 (3.90), *n*=7 | .00 [.00, 0.00], *n*=9* | 4.00 (5.00)* |
| Beck Depression Inventory-II Somatic Subscale | 2.82 (1.83) | 5.14 (4.41), *n*=7 | .00 [.00, 1.00], *n*=9 | 2.57 (2.07) |
| SF-36 Mental Health | 57.95 (4.33) | 46.84 (14.16), *n*=8 | 58.46 [55.64, 58.46] | 52.83 (9.34) |
| SF-36 Role Emotional | 55.88 [51.99, 55.88] | 53.94 [32.56, 55.88], *n*=8 | 55.88 [55.88, 55.88] | 55.88 [36.44, 55.88] |
| SF-36 Social Functioning | 56.85 [45.94, 56.85]* | 37.76 (15.97), *n*=8* | 56.85 [56.85, 56.85] | 45.94 [45.94, 56.85] |
| SF-36 Vitality | 51.24 (9.78) | 42.33 (15.87), *n*=8 | 58.33 [58.33, 61.46] | 52.09 (11.40) |
| SF-36 General Health | 53.00 (8.73) | 51.03 (7.81), *n*=8 | 57.23, [55.32, 62.47] | 52.46 (6.34) |
| SF-36 Bodily Pain | 52.17 (8.12) | 43.68 (15.33), *n*=8 | 55.36 [55.36, 55.36] | 53.49 (8.61) |
| SF-36 Role Physical | 49.28 (6.97) | 40.63 (15.54), *n*=8 | 56.85 [54.40, 56.85] | 50.55 (7.46) |
| SF-36 Physical Functioning | 51.77 [42.30, 57.03] | 52.82 [43.55, 55.98], *n*=8 | 54.93 [50.72, 57.03] | 54.93 [50.72, 57.03] |
| SF-36 Mental Component | 56.87 (5.41)* | 42.60 (15.44), *n*=8* | 56.90 [56.10, 58.19] | 49.69 (10.84) |
| SF-36 Physical Component | 47.80 (9.25) | 46.37 (11.02), *n*=8 | 56.16 [55.08, 57.48] | 53.30 (3.35) |
| Post-Lyme Questionnaire of Symptoms Total | 1.00 [.00, 1.00]* | 3.00 [1.00, 5.50], *n*=8* | .00 [.00, 1.00]* | 2.00 [1.00, 6.00]* |
| Post-Lyme Questionnaire of Symptoms Cognitive Subscale | 0.00 (0.00) | .00 [.00, 1.00], *n*=8 | .00 [.00, 1.00] | .00 [.00, .00] |
| Post-Lyme Questionnaire of Symptoms Neuro Subscale | .00 [.00, 1.00] | 1.75 (2.05), *n*=8 | .00 [.00, .00] | 1.00 [.00, 2.00] |
| Fatigue Severity Scale Total | 23.80 (12.44), *n*=10 | 34.13 (21.82), *n*=8 | 19.30 (9.37) | 22.71 (12.87) |
| Short Form McGill Pain Questionnaire Total | 1.00 [.00, 2.00], *n*=10 | 3.38 (4.17), *n*=8 | 0.50 [.00, 2.00] | 3.57 (4.43) |
| Short Form McGill Pain Questionnaire Affective Subscale | .00 [.00, 1.00], *n*=10 | .00 [.00, 1.50], *n*=8 | .00 [.00, .00] | 0.00 [0.00, 0.00] |
| Short Form McGill Pain Questionnaire Sensory Subscale | .50 [.00, 1.00], *n*=10 | 2.63 (3.29), *n*=8 | 0.50 [.00, 2.00] | 3.43 (4.12) |
| Mean (standard deviation) are presented for normally distributed continuous variables, and median [IQR] are presented for non-normally distributed continuous variables.<br>Sample *n* 's are presented for any measures with missing data.<br>Significant group differences are marked for each test * *p* = sPTLD vs. RTH, *p* < .05. |  |  |  |  |

### Cognitive assessment

At baseline, the HC group scored higher than the RTH and sPTLD groups on Trails A, (vs. RTH: *t(28) = 2.17, p = .038*; vs. sPTLD: *t(26) = 2.49, p = .019*). At follow-up, HC and patient groups showed no significant differences. The RTH vs. sPTLD groups showed no significant differences at either timepoint. (Online Table3)

Tests were run to identify changes in cognitive test performance between the two timepoints. A 2(timepoint: baseline vs. follow-up) by 3(group: HC vs. sPTLD vs. RTH) mixed-design ANOVA was run for each cognitive test that was administered. A main effect of timepoint was observed for the Digit Span test *(F(1, 30) = 4.76, p = .037)* and Trails A test *(F(1, 29) = 7.50, p = .010*), with all groups improving on both tests. There also was a main effect of group for the Trails A test *(F(1, 29) = 4.28, p = .024*). Post-hoc pairwise t-tests for Trails A indicated that the HC group’s improvement across visits was marginally significant, *t(15) = −2.04, p = .060*). By contrast, the patient groups showed no improvement across visits, (sPTLD: *t(5) = - 1.26, p = .263*; RTH: *t(9) = −1.53, p = .160)*.

### fMRI verbal working memory task

Mean accuracy and RT (for accurate trials only) were computed for the forward and control tasks at each timepoint. A 2(condition: control vs. forward) x 2(stimulus type: 1 vs. 2) x 3(group: HC vs. sPTLD vs. RTH) mixed-design ANOVA was conducted for the participants’ mean accuracy scores at the baseline visit. At baseline, there was an interaction of condition by stimulus type *(F(1, 35) = 7.78, p = .008),* which remained at follow-up *(F(1, 29) = 6.00, p = .021).* There were no significant effects of group on the mean accuracy at either timepoint. As shown in Online Table3, the condition by stimulus type interactions were due to reduced accuracy on the two-letters forward condition.

The same 2×2×3 mixed-design ANOVA was conducted for the mean RTs at each timepoint. At the baseline visit and at follow-up, there were interactions of condition by stimulus type *(F(1, 34) = 22.7, p < .001) and (F(1, 29) = 20.8, p < .001),* respectively. There were no significant effects of group on the mean RTs at either timepoint. As with the accuracy data, the condition by stimulus type interactions were due to increased RTs on the two-letters forward condition. Due to the disproportionate difficulty experienced by both groups on the two-letters forward condition, fMRI analyses focused on results obtained from this condition.

### MRI data

BOLD signal activations were compared between groups in a double subtraction approach: within-group contrast values for “two-letters forward” minus “two-letters control” conditions were obtained (first subtraction), then these values were compared between groups (second subtraction).

#### All Lyme vs. HC

At baseline, between-groups analysis revealed eight activations; all were in the direction of All Lyme participants showing elevated activation compared to HC. (Table 3) Strikingly, six of the eight activations (75%) were located in white matter, within the frontal lobe, temporal lobe, and cerebellum. With many areas of activation located in white matter, the observed brain circuitry did not align with prior reports using this fMRI task. [11, 22, 35] At 6-month follow-up, group differences revealed only two areas of activation, both in the direction of Lyme participants showing elevated activation compared to HC, and both located in the frontal lobe (one was in white matter).

**Table 3.** FMRI activation differences during the fMRI working memory task at baseline and 6-month follow-up.

|  |  |  |  |  |
| --- | --- | --- | --- | --- |
| **Cluster size (voxels)** | **T-value** | **X, Y, Z (MNI)** | **Brain Region (BA)** | **% White Matter** |
| **Baseline Regions of Activation** |  |  |  |  |
| **All Lyme > HC** |  |  |  |  |
| 67 | 4.31 | -42, -22, -8 | L Temporal Lobe | **91\*** |
| 36 | 4.25 | -16, -12, 36 | L Frontal Lobe | **100** |
| 65 | 4.04 | -36, -16, 54 | L Frontal Lobe | **56\*** |
| 54 | 4.03 | 12, -22, 68 | R Frontal superior gyrus (BA 6) | 47* |
| 26 | 3.83 | -32, 4, 36 | L Frontal Lobe | **95** |
| 31 | 3.68 | -16, -48, -28 | L Cerebellar Anterior Lobe | **97** |
| 11 | 3.66 | -50, 18, 18 | L Frontal Lobe | **61** |
| 21 | 3.58 | 2, -72, -36 | R Cerebellar Vermis VIIIB | 9 |
| **HC > All Lyme** |  |  |  |  |
| None |  |  |  |  |
| **RTH > HC** |  |  |  |  |
| 134 | 5.45 | -16, -12, 38 | L Frontal Lobe | **97\*** |
| 217 | 5.01 | -34, -18, 54 | L Frontal Lobe | **52\*** |
| 190 | 4.77 | 12, -22, 68 | R Superior frontal gyrus (BA 6) | 41* |
| 191 | 4.60 | 46, -8, 32 | R Precentral Gyrus (BA 6) | 41* |
| 63 | 4.33 | -42, -22, -8 | L Temporal Lobe | **87\*** |
| 198 | 4.19 | -30, 4, 36 | L Frontal Lobe | **78\*** |
| 152 | 4.00 | 30, -44, 24 | R Parietal Lobe | **99\*** |
| 64 | 3.98 | 2, -72, -32 | R Cerebellar Vermis VIIIB | 12* |
| 26 | 3.97 | -8, -20, 70 | L Frontal Lobe | **84** |
| 81 | 3.94 | 62, -8, 12 | R Postcentral Gyrus (BA 43) | 32* |
| 32 | 3.92 | -22, -56, 36 | L Parietal Lobe | **100** |
| 15 | 3.81 | -24, 22, 20 | L Frontal Lobe | **100** |
| 10 | 3.78 | 36, -10, -20 | R Hippocampus | 20 |
| 26 | 3.77 | -34, -36, 26 | L Parietal Lobe | **99** |
| **HC > RTH** |  |  |  |  |
| None |  |  |  |  |
| **sPTLD > HC** |  |  |  |  |
| 25 | 4.37 | -4, -40, -18 | L Cerebellar Lobule III | 29 |
| **HC > sPTLD** |  |  |  |  |
| None |  |  |  |  |
| **RTH > sPTLD** |  |  |  |  |
| 78 | 4.13 | 48, -10, 32 | R Post Central Gyrus (BA 4) | 9* |
| 10 | 3.93 | -18, -12, 40 | L Frontal Lobe | **100** |
| 34 | 3.93 | -52, 24, 8 | L Inferior Frontal Gyrus (BA 45) | 30<sup>^</sup> |
| 11 | 3.71 | -14, -60, 48 | L Parietal Lobe | **88<sup>^</sup>** |
| 36 | 3.58 | 38, -46, 20 | R Parietal Lobe | **95<sup>^</sup>** |
| 12 | 3.42 | 16, -4, 48 | R Frontal Lobe | **74<sup>^</sup>** |
| **sPTLD > RTH** |  |  |  |  |
| None |  |  |  |  |
| **6-Month Follow-up Regions of Activation** |  |  |  |  |
| **All Lyme > HC** |  |  |  |  |
| 32 | 4.38 | -28, 10, 60 | L Middle Frontal Gyrus (BA 6) | 4 |
| 10 | 4.02 | 34, 34, 6 | R Frontal Lobe | **96** |
| **HC > All Lyme** |  |  |  |  |
| None |  |  |  |  |
| **RTH > HC** |  |  |  |  |
| 17 | 4.60 | 34, 32, 6 | R Frontal Lobe | **78** |
| 10 | 3.86 | -28, 10, 58 | L Middle Frontal Gyrus (BA 6) | 1 |
| **HC > RTH** |  |  |  |  |
| None |  |  |  |  |
| **sPTLD > HC** |  |  |  |  |
| 50 | 5.81 | -30, 10, 62 | L Middle Frontal Gyrus (BA 6) | 8 |
| **HC > sPTLD** |  |  |  |  |
| None |  |  |  |  |
| **RTH > sPTLD** |  |  |  |  |
| 15 | 4.57 | 42, -80, 18 | R Occipital Lobe | **56** |
| 19 | 4.06 | 20, -64, 32 | R Precuneus (BA 7) | 7<sup>^</sup> |
| 52 | 3.80 | 44, -14, 36 | R Post Central Gyrus (BA 4) | 19<sup>^</sup> |
| 41 | 3.77 | 62, -4, 16 | R Post Central Gyrus (BA 4) | 16<sup>^</sup> |
| 10 | 3.62 | -14, -48, -40 | L Cerebellar Lobule IX | 48<sup>^</sup> |
| 21 | 3.58 | -24, -44, 34 | L Parietal Lobe | **88<sup>^</sup>** |
| **sPTLD > RTH** |  |  |  |  |
| 13 | 4.34 | -56, 20, 14 | L Inferior Frontal Gyrus (BA 45) | 0 |
| 17 | 4.30 | 58, 20, 20 | R Inferior Frontal Gyrus (BA 45) | 15 |
| Primary threshold = $p < .001$ and $k \geq 10$; <sup>^</sup> threshold = $p < .0025$ and $k \geq 10$.<br>MNI = Montreal Neurological Institute coordinates; BA = Brodmann Area.<br>Bold % white matter indicates > 50%.<br>For white matter > 50%, the brain region was reported within the relevant lobe.<br>* indicates the activation passed $p < .001$ and also surpassed $k \geq$ expected voxels per cluster (i.e., beyond $k \geq 10$ voxels) | | | | |

#### RTH vs. HC

At baseline, between-groups analysis yielded 14 activations, all in the direction of RTH showing elevated activation compared to HC. Of these, nine (64%) were in white matter. At follow-up, group differences revealed two activations, both in the direction of RTH showing elevated activation than HC and located in the frontal lobe (one was in white matter). (Fig. 2) Activation patterns at both time points closely resembled those observed in the All Lyme vs. HC comparisons, suggesting that the RTH group drove the All Lyme vs. HC results.

**Fig. 2.**
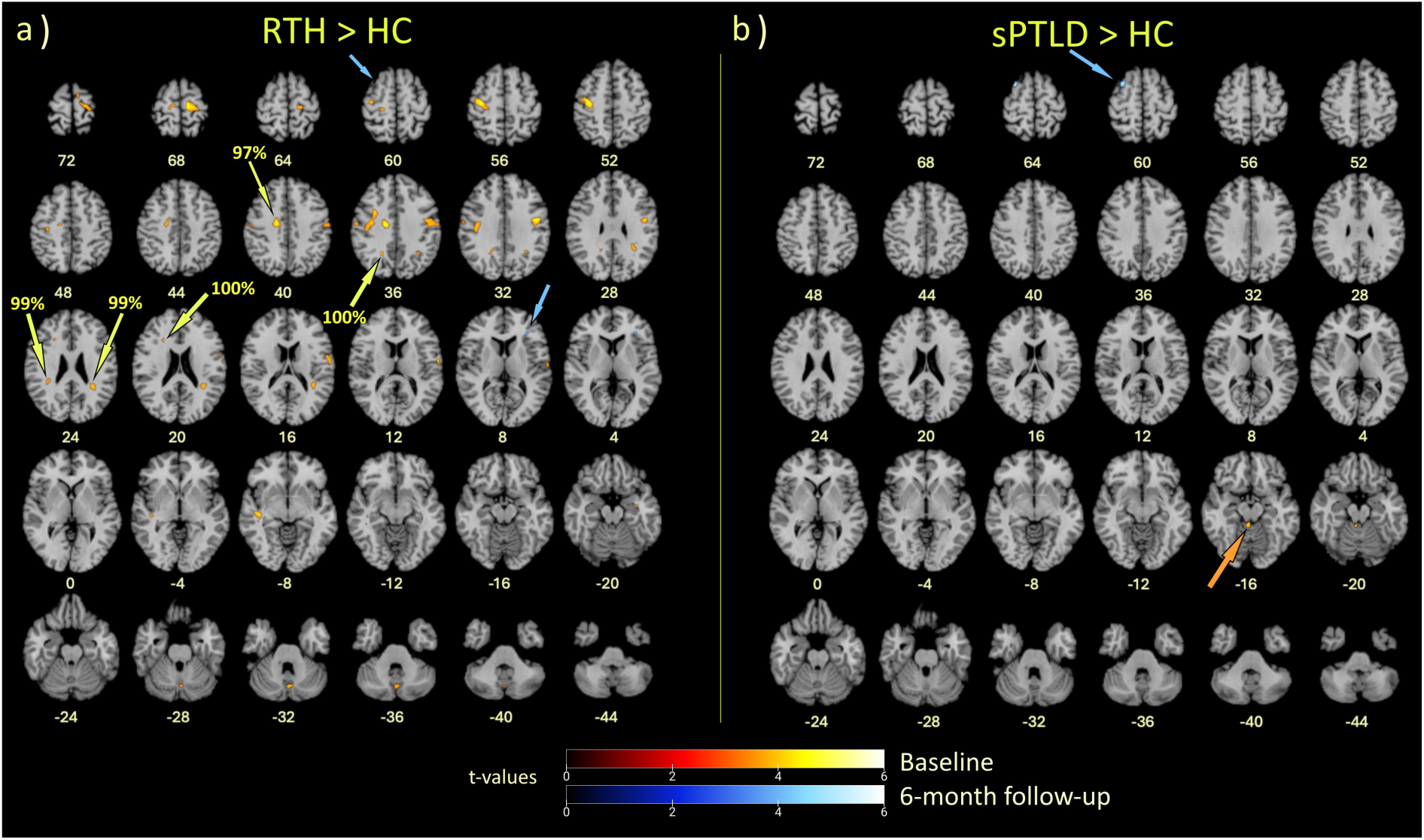
fMRI cluster overlays representing between-group activation differences during baseline and follow-up scans. **a)** RTH > HC: Numerous activation differences were evident at baseline (red-yellow spectrum clusters). Clusters that were > 90% white matter are denoted by the yellow arrows. Only two activation differences were observed six months later (blue spectrum clusters; blue arrows). **b)** sPTLD > HC: One activation difference at baseline and at 6-month follow-up were observed, indicated by orange and blue arrows, respectively. Cross-reference with Table 3 for cluster information. Scale shows t-values 0 – 6. Images are shown in neurological convention with right side on the right. Threshold = p < .001 and k ≥ 10.

#### sPTLD vs. HC

At baseline, between-groups analysis revealed one gray matter activation in the cerebellum, in the direction of sPTLD showing elevated activation compared to HC. At follow-up, group differences revealed one gray matter activation in the frontal lobe, reflecting elevated activation in sPTLD compared to HC. (Fig. 2)

#### RTH vs. sPTLD

At baseline, between-groups analysis revealed two activations, with RTH showing elevated activation compared to sPTLD. One activation was located in white matter, and both were in the frontal lobe. At follow-up, group differences revealed one area of elevated activation in occipital lobe white matter of RTH compared sPTLD. We expected to see more group differences in the direction of RTH stronger than sPTLD, given each subgroup’s results compared to HC. Therefore, we explored direct subgroup comparisons using a slightly more liberal threshold of p < .0025, k ≥ 10. Indeed, this revealed four additional activations at baseline, all with the RTH group showing stronger activations than the sPTLD group (three in white matter). At six months, five additional activations were revealed in favor of the RTH group showing stronger activations (one in white matter). (Table 3)

#### Longitudinal Comparisons

Within-group analyses compared activations at baseline and follow-up, but no suprathreshold clusters were revealed.

#### Tissue class segmentation results

At baseline, 75% of ROIs were located in white matter for All Lyme vs. HC. When RTH vs. HC was examined separately from sPTLD vs. HC, 64% of the ROIs were located in white matter, thereby driving the All Lyme vs. HC results. At the 6-month follow-up, there was a reduction in group brain activity differences, with the All Lyme vs. HC and RTH vs. HC comparisons nearly identical, each revealing two ROIs, with one of them in white matter. No white matter activations were observed in the sPTLD group at either timepoint.

#### Relationship between fMRI and clinical and cognitive scores

For clinical correlates, baseline activation signals from the ROIs in the RTH group generally correlated with better clinical outcomes, regardless of whether the ROIs were found in gray or white matter. (Fig. 3) Six months later, this relationship had reversed. ROI activity generally correlated with patients feeling worse in the RTH group. By contrast, in the sPTLD group at baseline, ROI activity generally associated with poorer clinical outcomes. Six months later, this relationship had dissipated without a clear association between brain activity and clinical outcomes.

**Fig. 3.**
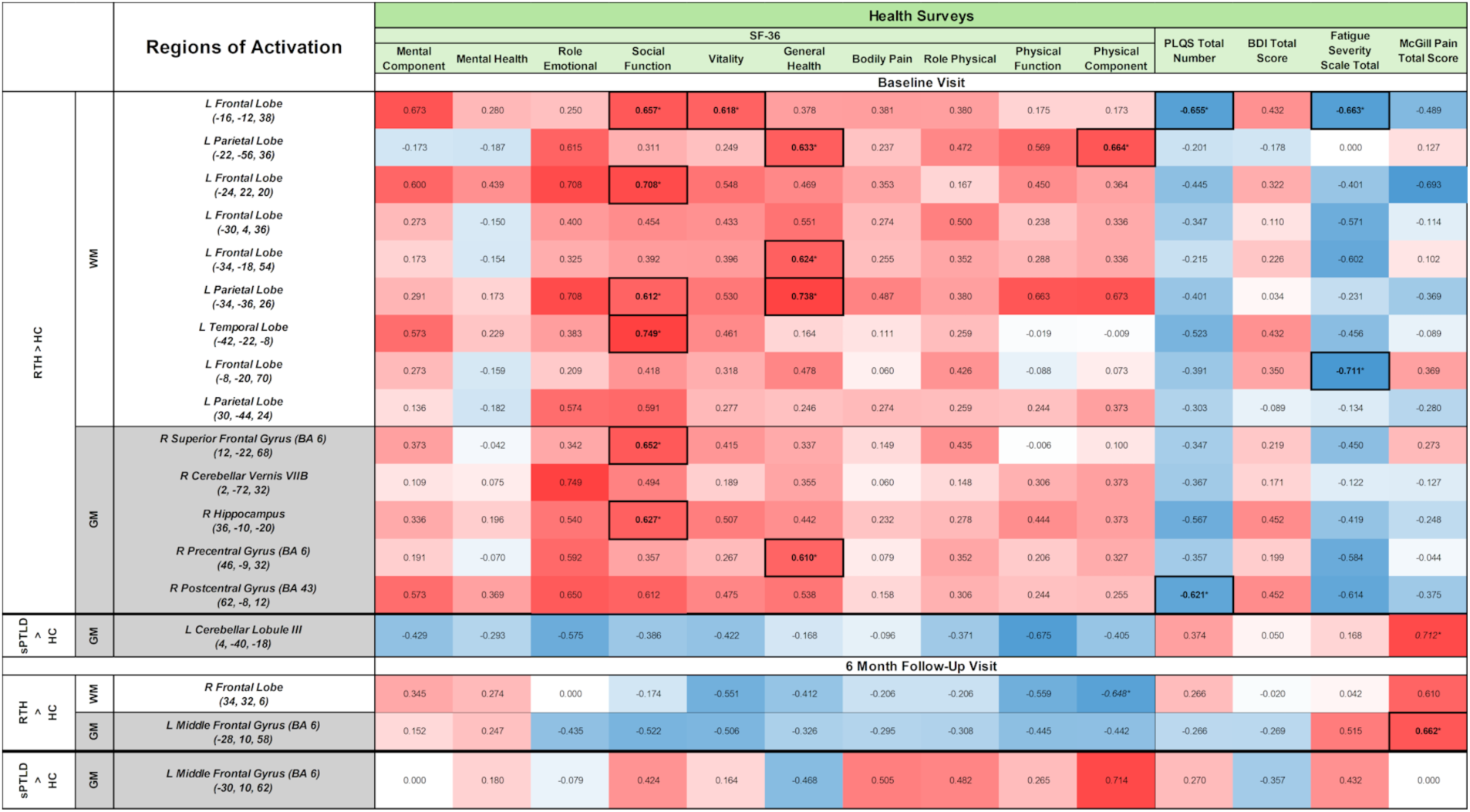
Correlation matrix for ROI values and clinical assessment scores. Shades of red = positive correlation direction; shades of blue = negative correlation direction. Note that the SF-36 uses an inverse relationship to severity of illness. Correlations were run using Spearman’s rank; significance is denoted by * <.05 if correlations also passed visual scatterplot inspection.

At the baseline visit, correlations between ROI values and cognitive scores did not show clear associations for either group. (Fig. 4) At follow-up, however, the sPTLD group skewed towards negative associations between brain activity and cognitive function, with significant correlates for assessments of processing speed and executive function (Digit Symbol Coding and Trails B). Negative correlates on additional tests (HVLT Total, Trails A, and COWA) were also quite strong, with *r_s_* > .6, which did not meet the threshold using non-parametric testing, but were consistent with difficulties in processing speed, attention, and language functions.

**Fig. 4.**
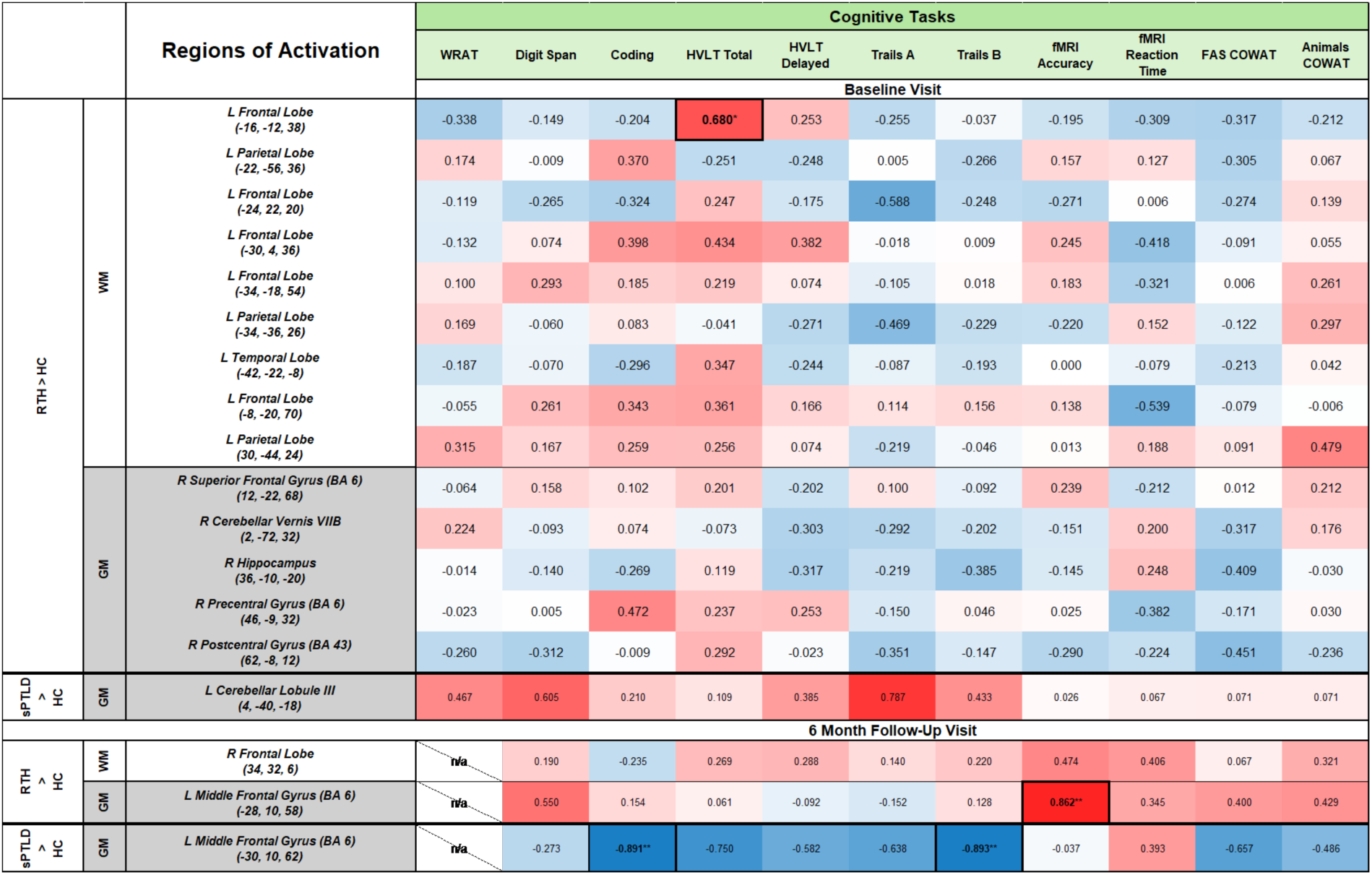
Correlation matrix for ROI values and cognitive test scores. Shades of red = positive correlation direction; shades of blue = negative correlation direction. Correlations were run using Spearman’s rank; significance is denoted by * <.05, ** < .01 if correlations also passed visual scatterplot inspection.

## Discussion

This study examined brain changes associated with early, antibiotic-treated Lyme disease. Early elevated fMRI, including in white matter, activity was associated with RTH status. Much of the observed activity was located in the frontal and parietal lobes, consistent with brain regions previously shown to be relevant to this working memory task in healthy participants. [11, 22, 24, 35] Moreover, the intensity of early fMRI activity correlated with better health scores, as indicated by self-report. Robust brain activity, in or out of white matter, was not observed in those who developed sPTLD. Thus, early brain activity does not appear to be a pathology. Rather, it seems to reflect an adaptive healing response that leads to a favorable prognosis.

Although there was no pattern of association between brain activity and health status in the sPTLD group, an association between brain activity and cognition was observed. Specifically, at the follow-up visit, elevated brain activity correlated with reduced cognitive performance for processing speed, executive function, and attention. This is consistent with reports in the literature of measurable cognitive impairments and subjective complaints of “brain fog” by PTLD patients.[7]

White matter activity during fMRI is unusual to observe due to this region’s low energy demands, which are about only about 25% that of gray matter. [13, 36–38] White matter includes myelin and axons that interconnect gray matter regions. Synapses are formed in gray matter and most of the energy (delivered by the blood) is needed for brain metabolism there, such as to enable action potentials, restore ion gradients, and release neurotransmitters. Thus, detection of white matter activity via fMRI methods is uncommon but has been reported in healthy adults. [13, 14, 39, 40] We were struck, however, by the large degree of white matter activity observed at baseline, and that this observation was specific to the RTH group. Such activity was mitigated, but not fully resolved, six months post-treatment. In the sPTLD group, white matter activity was not observed at all. Taken together, these findings suggest that white matter activity is an important aspect of an adaptive response to *Bb* infection and that early, vigorous white matter activity is a harbinger of a healthy outcome. Our prior findings showed that people who developed PTLD can also elicit white matter activity. [11] In that study, white matter activity also corresponded to better health reports, suggesting that even in the PTLD stage, the brain continues to try to heal.

However, questions remain: what does this white matter activity represent, how is it detected via fMRI, and why does it relate to symptom severity and recovery in LD? Several clues can be gleaned from the data observed here. First, the time course of white matter activity generally correlated with that of gray matter activity. This indicates that energy metabolism, measured by proxy using the BOLD signal, was required by both brain matter types in a temporally dependent, synchronous way. Second, because axons and myelin do not have sufficient vascularization to support a BOLD signal, the BOLD signal observed here actually may have originated from a source surrounding the axon, which could not be disentangled visually with fMRI spatial resolution. Third, this phenomenon appears to be non-pathological because it corresponds to better, not worse, health outcomes.

The underlying mechanism that leads to white matter BOLD detection in this scenario should adhere to these qualities. A promising candidate mechanism may be mediated by astrocytes. Astrocytic proliferation is a response to neuronal damage and can be helpful or harmful, depending upon the context and duration. [41–44] Astrocytes assist action potential propagation at the Nodes of Ranvier (NR) along the myelin sheath. [45] Thus, if myelin were damaged or the NR altered in the course of LD, this could increase astrocytic involvement in the white matter in an event-related (i.e., action potential) manner. This would correspond with concurrent astrocytic activity at the synapse in gray matter, where astrocytes use neurovascular coupling to help regulate blood flow needed for energy metabolism in an event-related manner. [46–48] These properties are consistent with the scenario of robust white matter activity correlating with healthy outcomes. Functional connectivity analyses in future studies could provide further insight into the interrelationship between white and gray matter networks.

An alternative interpretation is that neuroinflammation increased astrocytic metabolic demand, leading to secondary white matter BOLD activity that was neither adaptive nor compensatory. [49] This, too, could explain the mitigated response at six months after healing had occurred. However, if so, it is unclear why a similar pattern of white matter BOLD activity was not observed at either timepoint in the sPTLD group who presumably also experienced neuroinflammation.

While astrocytic involvement is plausible, this needs to be confirmed via direct astrocytic markers. Astrocytic reactivity can be measured in the cerebral spinal fluid (CSF) and blood serum by the protein glial fibrillary acidic protein (GFAP). Although GFAP levels were not obtained in the current study, increased GFAP levels were reported in patients with untreated LD long ago. [50] Notably, increases were detected within weeks of infection, decreased 4-8 weeks after antibiotic treatment, and correlated with duration of symptoms. This, too, fits with our pattern of findings. Future studies should examine the phenomenon of white matter activity in LD more pointedly, including additional measures that target white matter integrity (e.g., diffusion tensor methods), in combination with GFAP and other biomarkers, to better understand how brain activity relates to health outcomes.

White matter BOLD signal could also result from head motion or physiological noise during fMRI acquisition. In this study, image processing included motion correction to reduce this possibility. It is unlikely, however, that head motion would result in white matter BOLD signal revealed squarely within white matter tracts, in many cases > 90%, rather than along the edges of the brain or ventricle surfaces, as is often the case with head motion. It is also highly unlikely that motion artifacts occurred within one group at one timepoint only, with all other variables remaining constant (e.g., MRI scanner, scan sequence, analytic methods, etc.), and that the MRI values derived from these regions correlated with patient-reported health outcome scores, the latter being a replication of our prior findings in PTLD. [11] Nonetheless, white matter activity is an unusual finding and deserves to be interpreted with caution.

Dividing the LD group resulted in relatively small sample sizes for each subgroup. However, the robustness of activity observed in the RTH vs. HC comparison with only n=11 in the RTH group is impressive at p < .001. Small sample sizes likely masked group differences in direct comparison between the two LD subgroups, which were revealed when a slightly more liberal threshold of p < .0025 was applied. Larger-scale longitudinal studies are needed to confirm these findings. Because brain activations observed in the RTH group were reduced, but not entirely absent, by six months, participants should be followed longer than six months to identify if full resolution occurs.

In summary, this study demonstrated that an indicator of RTH status after *Bb* infection and treatment was early, robust brain activity located primarily in white matter. We postulate the source of white matter activity is related to astrocyte function, but this requires further investigation. Understanding how elevated white matter activity is related to RTH in those infected by *Bb* could aid early identification of those most vulnerable to developing PTLD and guide treatment.

## Supporting information

Supp_Methods1 fMRI parameters

Supp_Methods2 MRI equipment

Supp_Table1 clinical assessment

Supp_Table2 cognitive assessment

Supp_Table3 cognitive results

## Data Availability

All data produced in the present study are available upon reasonable request to the authors

## Acknowledgments

We thank Jason Creighton, Susan Joseph, and Cheryl Novak for their assistance with recruitment, screening, and data collection.

## Author Contributions

The following authors contributed to the study conceptualization and design: Cherie Marvel, Alison, Rebman, Kylie Alm, Pegah Touradji, Arnold Bakker, Erica Kozero, Arun Venkatesan, Abhay Moghekar, and John Aucott. Data collection, curation, and analysis were performed by: Cherie Marvel, Alison Rebman, Kylie Alm, Pegah Touradji, Prianca Nadkarni, Deeya Bhattacharya, Owen Morgan, Amy Mistri, Erica Kozero, and Ashar Keeys. The first draft of the manuscript was written by Cherie Marvel and all authors commented on previous versions of the manuscript. All authors read and approved the final manuscript.

## Statements and Declarations

### Ethics approval

This study was performed in line with the principles of the Declaration of Helsinki. Approval was granted by the Institutional Review Board of the Johns Hopkins University School of Medicine (IRB# 00212903).

### Consent to participate

Informed consent was obtained from all individual participants included in the study.

### Data availability

De-identified data not published within this article may be shared at the request of any qualified investigator for purposes of replicating procedures and results contingent upon approval of a data sharing agreement.

