## Supplementary material for "Early brain changes in Lyme disease are associated with clinical outcomes": Supp_Methods1 fMRI parameters

**Supplement Methods1**

Additional fMRI verbal working memory task parameters

Participants were given up to 6 seconds to respond by button press (match = right index finger; non-match = right middle finger). Trials were jittered with an inter-trial interval (ITI) of 6-9 seconds. Response time (RT) and accuracy were recorded for each trial. Each participant completed one block each of the control and forward conditions, in separate scanner runs, with the order counterbalanced across participants. Each block contained 64 trials (~ 16 minutes). Probe letters matched a target (or newly derived target) on 50% of the trials. Parameters were pseudorandomized such that identical presentation occurred in no more than three consecutive trials: number of target letters (one or two), rehearsal duration (4 or 6 seconds), expected response (match or non-match), and duration of ITI (6–9 seconds).
