## Supplementary material for "Early brain changes in Lyme disease are associated with clinical outcomes": Supp_Methods2 MRI equipment

**Supplement Methods2**

MRI equipment information

Stimuli were delivered using E-Prime 2.0 software (Psychology Software Tools, Pittsburgh, PA) on a Dell Inspiron 7472 laptop running Windows 10 Pro. Participants viewed stimuli via an Epson PowerLite 7600p projected that projected onto a screen in the MRI scanner bore, which was then reflected onto a mirror attached to the top of the head coil and inclined at 45°. Due to an equipment upgrade during the study, one Lyme participant at baseline and three Lyme participants at the 6-month follow-up visit viewed stimuli via a Cambridge Research Systems BOLDscreen 32 UHD LCD display, which was run through a mirror box, then projected onto the head coil-mounted mirror. Button-press responses were collected using two fiber optic button boxes (MRA, Inc., Washington, PA) held by the participant in their right hand during the tasks.
