## Supplementary material for "Early brain changes in Lyme disease are associated with clinical outcomes": Supp_Table1 clinical assessment

### Supplement Table1. Clinical Assessment

| Questionnaire | Function | Scoring |
|---|---|---|
| Short Form Health Survey, version 2 (SF-36)[1] | 36-item measure of functioning in eight health attributes: Physical Functioning, Role of Physical (on daily living), Bodily Pain, General Health, Vitality, Social Functioning, Role of Emotional (on daily living), and Mental Health | Scores can also be compared to the US population mean (50.0 ± 10.0) |
| Fatigue Severity Scale (FSS) | 9-item measure of the impact of fatigue on daily function | Total scores ranging from 9 - 63 |
| Short-Form McGill Pain Questionnaire-2 (SF-MPQ-2) | 15-item pain metric | Total scores ranging from 0 - 45 |
| Beck Depression Inventory II (BDI-II) | 21-item depression metric | Total scores ranging from 0 – 63 with the following clinical classifications: 0-13, minimal depression; 14-19, mild depression; 20-28, moderate depression, and 29-63, severe depression |
| Post-Lyme Questionnaire of Symptoms [PLQS] | 36-item measure of symptom presence and severity | Total scores range from 0 - 36. |
| For all questionnaires, higher scores indicate more severe symptoms, except on the SF-36 where higher scores indicate higher health-related quality of life and functioning. | | |
