## Supplementary material for "Early brain changes in Lyme disease are associated with clinical outcomes": Supp_Table2 cognitive assessment

### Supplement Table2. Cognitive Assessment

| Standardized Test | Measurement |
| --- | --- |
| Wide Range Achievement Test – 4th Edition (WRAT-4)[1] | pre-morbid intellectual functioning |
| Digit Span (DS) Subtest of the Wechsler Adult Intelligence Scale – 4th Edition (WAIS-IV)[2] | attention (DS Forward) and working memory (DS Backward and DS Sequencing) |
| Hopkins Verbal Learning Test – Revised (HVLT-R)[3] | verbal list learning, delayed recall and retention, and recognition memory |
| Trail Making Test (Trails A and Trails B)[4] | processing speed, visual scanning, attention, and psychomotor processing; Trails B additionally included executive function |
| WAIS-IV Digit Symbol Coding[2] | processing speed and working memory |
| Controlled Oral Word Association Test (COWA)[5] | verbal fluency, using letter and semantic cues under time constraints |
