## Supplementary material for "Early brain changes in Lyme disease are associated with clinical outcomes": Supp_Table3 cognitive results

| **Supplement Table3. Cognitive assessment summary data** | | | | | | |
|---|---|---|---|---|---|---|
| | **Baseline Visit** | | | **6-Month Follow-up** | | |
| | RTH (*n*=11) | sPTLD (*n*=9) | HC (*n*=19) | RTH (*n*=10) | sPTLD (*n*=7) | HC (*n*=16) |
| Wide Range Achievement Test | 61.64 (7.55) | 61.67 (11.65) | 53.00 [50.00, 63.00], n=17 | N/A | N/A | N/A |
| Digit Span | 50.91 (9.30) | 49.67 (8.54) | 51.32 (9.62) | 51.40 (11.24) | 51.43 (7.55) | 57.69 (13.31) |
| Digit Symbol Coding | 54.73 (8.30) | 53.67 (7.28) | 60.00 [53.00, 63.00] | 58.00 (6.93), n=10 | 54.42 (6.43), n=7 | 61.06 (8.17) |
| Hopkins Verbal Learning Total | 50.09 (7.49) | 51.11 (11.14) | 55.0 (6.77) | 51.30 (8.91), n=10 | 56.43 (13.60), n=7 | 58.19 (9.22) |
| Hopkins Verbal Learning Delayed Recall | 53.45 (8.47) | 50.67 (11.53) | 52.16 (7.74) | 52.00 (8.25), n=10 | 56.71 (6.69), n=7 | 60.00 [51.25, 61.00] |
| Hopkins Verbal Learning Retention | 53.64 (7.62) | 53.00 [46.50, 57.00] | 49.32 (7.70) | 52.6 (6.57), n=10 | 56.29 (5.31), n=7 | 53.44 (6.52) |
| Hopkins Verbal Learning Recognition | 54.00 [51.00, 58.00] | 58.00 [52.00, 58.00] | 51.00 [51.00, 58.00] | 49.40 (7.06), n=10 | 57.50 [37.00, 58.25], n=6 | 58.00 [50.25, 58.00] |
| Trails A[BL: #^&; 6M: #] | 47.55 (5.77) | 43.78 (10.24) | 53.63 (9.56) | 53.20 (9.26), n=10 | 49.00 [38.00, 56.00], n=6 | 58.50 [51.50, 65.75] |
| Trails B | 51.73 (6.94) | 51.56 (11.10) | 55.00 (10.35) | 56.50 (5.94), n=10 | 54.14 (11.07), n=7 | 58.5 (11.80) |
| COWA verbal letters | 47.40 (11.34), n=10 | 48.00 (9.95), n=7 | Not Done | 51.14 (9.56), n=7 | 52.00 (5.55), n=6 | Not Done |
| COWA verbal animals | 52.20 (9.65), n=10 | 49.86 (13.33), n=7 | Not Done | 56.29 (12.08), n=7 | 63.50 [48.25, 68.25], n=6 | Not Done |
| <u>fMRI Task</u><br>Accuracy Control Condition 1 Stimulus | 100.00 [93.80, 100.00 | 96.89 (3.13) | 96.90 [96.90, 100.00] | 98.45 [95.33, 100.00], n=10 | 100.00 [96.90, 100.00], n=7 | 96.90 [93.80, 100.00], n=15 |
| Accuracy Control Condition 2 Stimuli | 93.80 [93.90, 100.00] | 100.00 [80.05, 100.00] | 96.90 [93.80, 100.00] | 96.90 [93.00, 100.00], n=10 | 100.00 [93.80, 100.00], n=7 | 96.90 [90.60, 100.00], n=15 |
| Accuracy Forward Condition 1 Stimulus | 95.35 [87.50, 100.00], n=10 | 100.00 [93.75, 100.00] | 96.90 [93.80, 100.00] | 95.96 (3.90), n=10 | 96.90 [96.90, 100.00], n=7 | 100.00 [96.90, 100.00], n=15 |
| Accuracy Forward Condition 2 Stimuli | 89.37 (5.56), n=10 | 91.69 (9.75) | 91.63 (4.79) | 89.70 (7.95), n=10 | 91.07 (6.09), n=7 | 93.80 [87.50, 96.90], n=15 |
| Response Time Control Condition 1 Stimulus | 918.84 (163.52) | 799.38 (130.16), n=8 | 912.36 (184.64) | 912.51 [676.48, 963.68], n=10 | 829.13 [632.17, 861.88], n=7 | 878.61 [775.59, 1015.26], n=15 |
| Response Time Control Condition 2 Stimuli | 1065.93 (187.27) | 913.04 (98.73), n=8 | 1006.21 [870.15, 1100.32] | 932.71 (157.18), n=10 | 886.54 (144.62), n=7 | 1043.43 (233.12), n=15 |
| Response Time Forward Condition 1 Stimulus | 959.70 (200.50), n=10 | 881.71 (151.39) | 1000.25 (211.90) | 869.69 (126.45), n=10 | 756.89 (114.13), n=7 | 972.77 (206.48), n=15 |
| Response Time Forward Condition 2 Stimuli | 1256.09 [1057.77, 1898.28], n=10 | 1201.30 (245.18) | 1368.65 (373.91) | 1175.62 (263.29), n=10 | 1038.04 (184.73), n=7 | 1176.45 [932.78, 1515.86], n=15 |

Mean (standard deviation) for normally distributed continuous variables, Median [IQR] for non-normally distributed continuous variables, and sample n's are presented for any measures with missing data.

Significant group differences are marked for each test, $p < .05$

BL = Baseline; 6M = 6-month follow-up; '*' = RTH vs. sPTLD; '#' = All Lyme vs. HC; '^' = RTH vs. HC; '&' = sPTLD vs. HC
